# Machine Learning Model to Predict Allocation of Patients with Chronic Back Pain for Integrated Practice Units in a System of Value-based Health Care

**DOI:** 10.1101/2023.11.05.23298111

**Authors:** Vitor Pereira Barbosa, João Lucas Maehara Said dos Reis, Cyro von Zuben de Valega Negrão, Victor Rossetto Barboza, Keli Cristina Betto Simoes Marcondes, Elton Pereira Rezende, Natália Neto Pereira Cerize, Vinicius Monteiro de Paula Guirado

## Abstract

Chronic pain incurs substantial global healthcare costs, requiring long-term specialized care. However, the variability in treatment approaches hampers the feasibility of value-based healthcare. To advance management and prevention efforts, data science tools like supervised and unsupervised machine learning algorithms offer promising solutions. This study employed data from six questionnaires to evaluate pain conditions in patients. Correlation techniques were applied to determine the questions most strongly correlated with low back pain, back pain, and leg pain. Five machine learning algorithms predicted the presence or absence of pain in the low back region. Seven variables were identified as input for the prediction models, with the XBoost Classifier demonstrating the highest accuracy, precision, recall, and F1-Score (0.8 accuracy, precision, recall; 0.78 F1-Score). By incorporating the machine learning model, it predicts the allocation of chronic back pain patients to integrated practice in a value-based healthcare system. Finally, our findings show that data science methods, such as machine learning, improve chronic pain management, expedite surveys, and improve value-based healthcare through efficient allocation.

**Graphical abstract:** 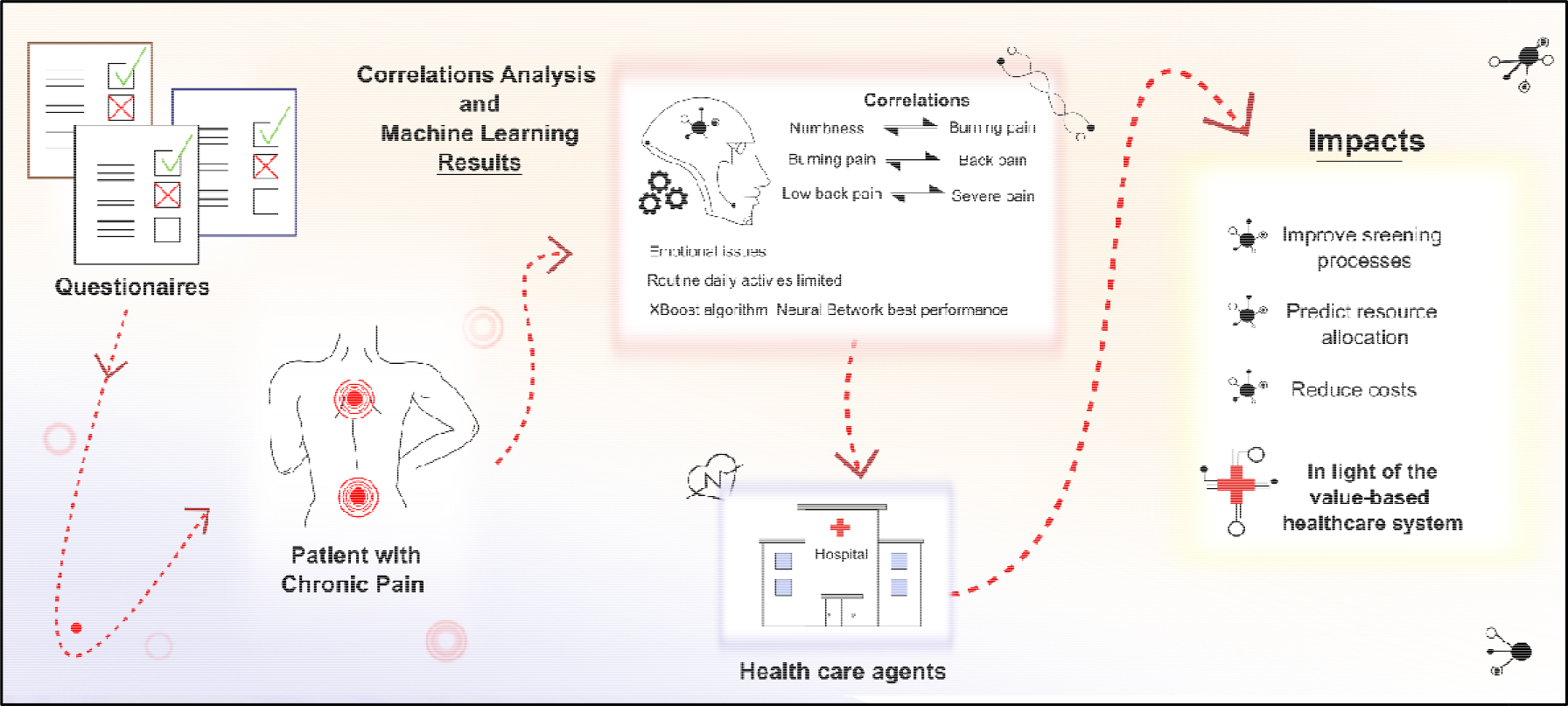

**Highlights:**

- Correlation analysis identified pain-related questions (back, leg, chronic low back).
- 7 characteristics used by ML algorithms to forecast chronic low back pain occurrence.
- XBoost algorithm and neural network achieved best performance (Precision, Recall ∼0.8).
- Outcomes aid patient allocation, screening, and reducing costs in value-based healthcare.
- Streamlining surveys boosts response rates and enables larger datasets for improved modeling.

## 1. Introduction

One of the primordial questions proposed by Porter et al. in the book “*Redefining Health Care*” was why the competitive management model failed the healthcare system ^1^. The author argues, for example, that throughout history, the economy and competition in the private market have been one of the most significant forces for the improvement of quality and cost reduction in goods and services. However, this was not observed in the health sector, where the competition only acted on the costs, which grew, and the quality of the services did not necessarily improve ^1^.

The necessity of a value-based healthcare system arose from the high and unsustainable costs of the current practiced system ^2^. This analysis was initially carried through the American Healthcare System, which presented a fundamental paradox related to the increase in biomedicine knowledge, which was the protagonist in innovations in therapies and surgical procedures and the treatment of previously fatal conditions ^3^. However, this system started to present problems in basic questions related to quality, results achieved for the patients, and costs ^3^.

In 2006, the *National Academy of Medicine* established the basis for evidence-based medicine to provide a reliable base for national healthcare leaders to allow the generation of a system that can generate real value for the patients and society. The purpose of advancing up to one *Learning Health System* quickly emerged and was defined as a system where science, computer science, incentives, and culture are lined up for improvement and continuous innovation ^4^.

Considering the global scenario, the problems related to the increase of costs are also cited in a report elaborated by Deloitte company in 2019, which pointed out that the global expenditure in healthcare was expected to grow to an annual tax of 5.4% between 2018-2022, compared with an increase of 2.9% between 2013-2017 ^5^. This estimate was based mainly on the strengthening of the dollar when compared to the euro and other currencies; the expansion of the coverage of medical assistance in developing countries; the aging population; the sprouting of new treatments and technologies in healthcare, and the increase of labor costs in the health sector ^6^.

In the specific field of neurological surgery, diagnostic and therapeutical options are available in a highly complex scenario at the beginning of the 21st century ^7^. Likewise, the range of possible results is varied because many dimensions of interpretation exist. Still influenced by the social context, patients and doctors are overwhelmed with information from the digital age, and thus, the decision-making process today is cardinal and critical^7,8^. The contemporary resource for this challenging demand is the application of information management technologies, such as artificial intelligence ^9^.

Recent advances in artificial intelligence (AI) create new opportunities to personalize technology-based health interventions for patients^9^, including for chronic pain^10,11^. Intelligence learning environments, interaction narrative generation, user modeling, and adaptative training tools available in AI model the learning and involvement of patients with chronic pain and provide personalized support in adaptative health technologies ^10^. Many of these technologies have emerged from applications centered on human activities for education, training, and entertainment. However, its application in health improvement, so far, has been comparatively limited.

Chronic spinal pain is part of the chronic pain category, which affects approximately 20 % of the world’s population^12,13^. Primary chronic pain is defined as pain that persists for more than three months and has a significant impact on emotional welfare, being a definite cause of distress, demoralization, and functional disability in patients, which makes it one of the primary sources of suffering ^14^.

There are two well-established pain classification systems, *STarT Back* and *McKenzie*. The *McKenzie* method uses the patient’s symptom history and pain presented after conducting specific movements and classifies them into three groups according to their syndrome. Based on physical and psychosocial factors, the *STarT Back* classifies the patients as having high, medium, and low risk of developing persistent symptoms that disable them. These two methods are examples of approaches that do not consider only the anatomical basis to perform the diagnosis ^15^. However, these methods have disadvantages, as shown by Tagliaferri, with highly varied results related to reliability, construct validity, and discriminative validity concerning Machine Learning tools ^16^.

The presented scenario highlights the inadequacy of oversimplifying complexity as a solution in neurological surgery. Instead, integrating science with data, combining human knowledge and digital technologies, emerges as the optimal alternative for informed decision-making. By embracing digital technologies, healthcare professionals gain the opportunity to enhance and streamline the clinical routine, as well as improve patient management through data-supported interventions. This utilization of digital tools facilitates the decision-making process and paves the way for more effective and efficient healthcare delivery in neurological surgery.

This study uses exploratory data analysis, pre-processing techniques, and machine learning models on the available database to find the essential variables that display the most vital link with chronic low back pain, back pain, and leg pain. Using these techniques, we want to identify the elements most closely related to these kinds of pain, enabling a more profound comprehension of their underlying origins. The potential for AI-driven adaptive solutions in preventative healthcare for people with chronic pain is also highlighted by this research. We forecast how future treatments for this patient population might be carried out inside and outside specialized clinics. We foresee a proactive approach to healthcare that focuses on preventing illness by leveraging the potential of artificial intelligence.

## 2. Materials and Methods

### 2.1. Data collection

The available data are the clinical evaluation results in a multidisciplinary integrated care unit specialized in chronic pain from February to December 2019. The initial available sample included 240 patients undergoing clinical evaluation. The data from the questionnaires are of a socio-demographic and clinical nature, which are listed below:

1. Basic clinical assessment of spine symptoms (Gothenburg Protocol);
2. Brief Pain Inventory (BPI);
3. Oswestry 2.0 Disability Index;
4. Roland Morris Disability Questionnaire (RMDQ);
5. Questionnaire to assess the quality of life 12-Item Short Form Health Survey (SF-12);
6. Questionnaire for Diagnosis of Neuropathic Pain 4 (DN-4).

### 2.2. Data pre-processing

The questions in the questionnaires were divided into two categories: nominal (all converted to binary format) and ordinal. This approach allows the correlation between the binary variables to be observed through a contingency table and the correlation between binary and ordinal variables to be measured with the Point-Biserial Correlation Coefficient.

The initial database results from a six questionnaire junction and comprises 118 variables (also mentioned as characteristics or column vectors). The dummy encoding technique was used to generate new columns for the variables presented as nominal.

As a consequence of this first pre-processing measure, 146 column vectors were obtained, among which 101 are binary and 45 are ordinal.

For applying the machine learning model for patient classification regarding the presence or absence of low back pain, patients who did not fill this question, corresponding to the pain of number 30 in the Brief Pain Inventory questionnaire, were removed from the dataset. As a result, there were 138 patients in the final database. This dataset can, therefore, be submitted to correlation analysis and graph construction and used as input for machine learning algorithms.

It is known that with this amount of column vectors (146) and only 138 patients, there is a case of sparse data in which Bellman (1957) introduced the phenomenon of the Curse of Dimensionality. The sparse data becomes a problem in obtaining results with statistical significance in a machine learning model because the number of observations should grow exponentially with the dimension (number of variables or factors) ^18^.

To reduce the number of predictor variables, we used the results derived from the analysis of the correlation between binary variables and the correlation between binary variables and ordinal variables to select those with the highest absolute values.

Additionally, as a tool that allows us to see the group’s division, the PCA analysis with two components was used to visualize the data belonging to the group of people who present low back pain and those who answered that they do not present this pain.

#### 2.2.1 Correlations and Statistics Model validation

This section describes the methods for correlating binary variables and correlating binary and ordinal variables, which were used for variable selection, also named feature selection.

##### Correlation between binary variables

To evaluate the correlation between binary variables, a contingency table was used, with its structure presented in Table 1 :

**Table 1.** Example of cross table.

|  |  | Variable B |  |
| --- | --- | --- | --- |
|  |  | 0 (not) | 1 (yes) |
| Variable A | 0 (not) | a | b |
|  | 1 (yes) | c | d |

Table 1 shows an example of a contingency table in which *a, b, c*, and *d* are integers. This table structure makes it possible to assess whether the change in proportion between no / yes answers of variable A is correlated with variable B.

To assess the statistical relevance of the results, the chi-square hypothesis test was applied:

H0 Independence - A does not depend on B;

H1 Dependence - A depends on B.

The chi-square calculation is done using the following expression:

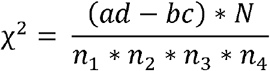

In which:

*a, b, c*, and *d*, are the observation counts presented in Table 1

*n*_1_ : *a* + *b*

*n*_2_: *c* + *d*

*n*_3_: *a* + *c*

*n*_4_: *b* + *d*

*N*: *n*_1_ + *n*_2_ + *n*_3_ + *n*_4_

This hypothesis test can be interpreted as variable A’s frequency distribution difference due to variable B’s presence. In such a manner, when the calculated chi-square value is greater than or equal to the tabulated chi-square when adopting a significance level of 5%, one can reject the null hypothesis of independence and consider the alternative hypothesis of dependence on variable A concerning variable B ^19^.

##### Correlation between binary and ordinal questions

The correlation between questions appears to be dichotomous, and questions that are present on an ordinal scale were performed through the Point-Biserial Correlation Coefficient.

It was performed a division in two groups: (0 - no) and (1 - yes); therefore, the coefficient can be calculated according to the following expression:

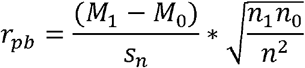

In which:

*s*_*n*_ : Standard deviation considering all population data.

*M*_0_ and *M*_1_: Average of the ordinal variables for patients who answered the question as negative (0 - no) and positive (1 - yes), respectively.

*n*_0_ and *n*_1_: Number of people in each group (0 - no) and (1 - yes).

The statistical significance of this coefficient is evaluated by the hypothesis test of Pearson’s correlation on the assumption that the biserial point correlation is a specific case for one of the dichotomous variables. The hypotheses are stated as follows:

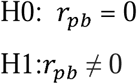

The test makes use of the *t* of *student* distribution so that the calculated *t value* is given by:

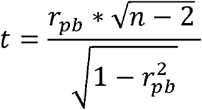

In which:

*r*_*pb*_: Biserial Point Correlation Coefficient.

*n*: Number of observations.

According to the adopted statistical criteria, if the value of calculate *t* is higher than the value of *t* tabulated when adopting a significance level of 5 %, the null hypothesis can be rejected ^20^.

Thus, only the correlations that passed this test were presented in this paper.

### 2.2. Sampling

The sampling technique used was K stratified folds (*StratifiedKFold*), with ten subdivisions (K = 10), to maintain the percentage of the original sample base for each class.

#### Supervised machine learning model

The performances of the following supervised machine learning models for classification were evaluated:

- Logistic regression with l1 and l2 regularization (*elasticnet*), with regularization factor l1 of 0.8. The optimization algorithm used to obtain the coefficients was lbfgs (*Limited-memory Broyden – Fletcher – Goldfarb – Shanno*);
- Neural network with one hidden layer composed of 5 neurons with the *ReLu* (rectified linear unit) activation function and on the output layer with the sigmoid activation function. The network was trained through discrete sample sizes of 5 observations for 200 epochs;
- *Random Forest:* 50 decision trees with an average depth of 8 levels were used. The criterion chosen to perform the split was *gini impurity;*
- SVM (*Suppor Vector Machine*): The Radial Base Function kernel, through the scalar product tools and the expansion of the Taylor series, was used to obtain a relationship between the observations in an infinite dimension. The probabilities generated by this model were calibrated using the Platt scaling calibration method, which applies the logistic regression function over the original generated probabilities, as detailed in ^21^;
- XBGClassifier: Implementation of the *Gradient Boosting* method with greater speed and design. It uses the so-called weak learners, decision trees with only one node and two leaves, so that through the method of the ensemble (joining of several models) can obtain an optimized and robust model to be used in classification in this study ^22^.

#### Model evaluation metrics

In order to evaluate the model’s performance on the correct classification of patients regarding the presence of chronic low back pain, the metrics Accuracy, *Precision, Recall*, and *F1-Score* were employed within the cross-validation process.

These metrics can be calculated from the confusion matrix, with its structure in Table 2.

**Table 2.**
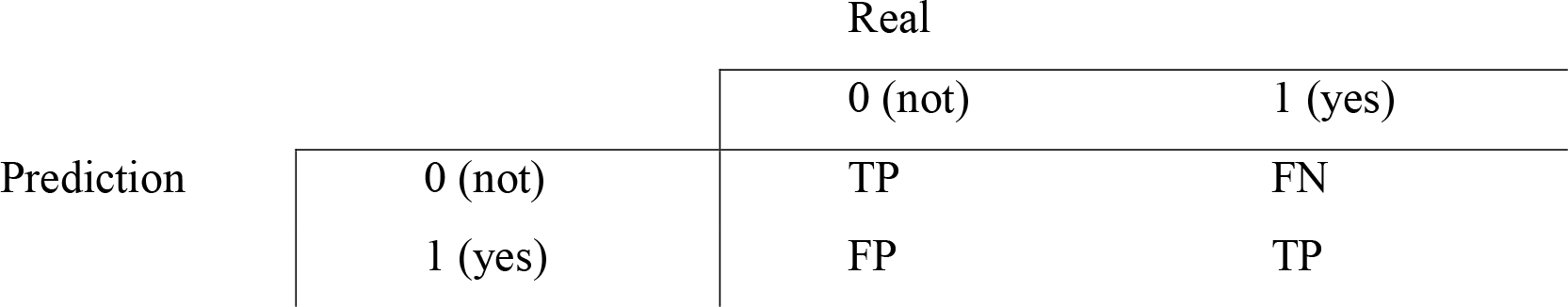
Example of the confusion matrix.

Based on what was exposed, the following metrics are defined:

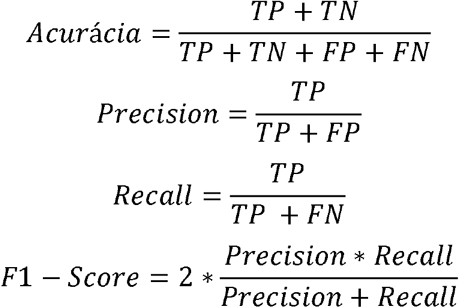

The metric *precision* calculates the ratio between the number of correctly predicted positives and all predicted positives. However, the Recall metric is critical considering this analysis’s focus, given that the calculation consists of the ratio between the total value of True Positives and the sum of True Positives and False Negatives ^23^.

The reason for using it is that a machine learning model for predicting the occurrence or not of chronic low back pain in patients must have a low amount of False Negatives and consequently a high *Recall* value. In addition, the F1-Score metric allows us to obtain a harmonic average between *Precision* and *Recall* ^24^.

The evaluation of the model was also carried out using curves ROC curve and AUC (*Area Under Curve*) Figure 1 (a) and *Precision-Recall curve* Figure 1 (b).

**Figure 1.**
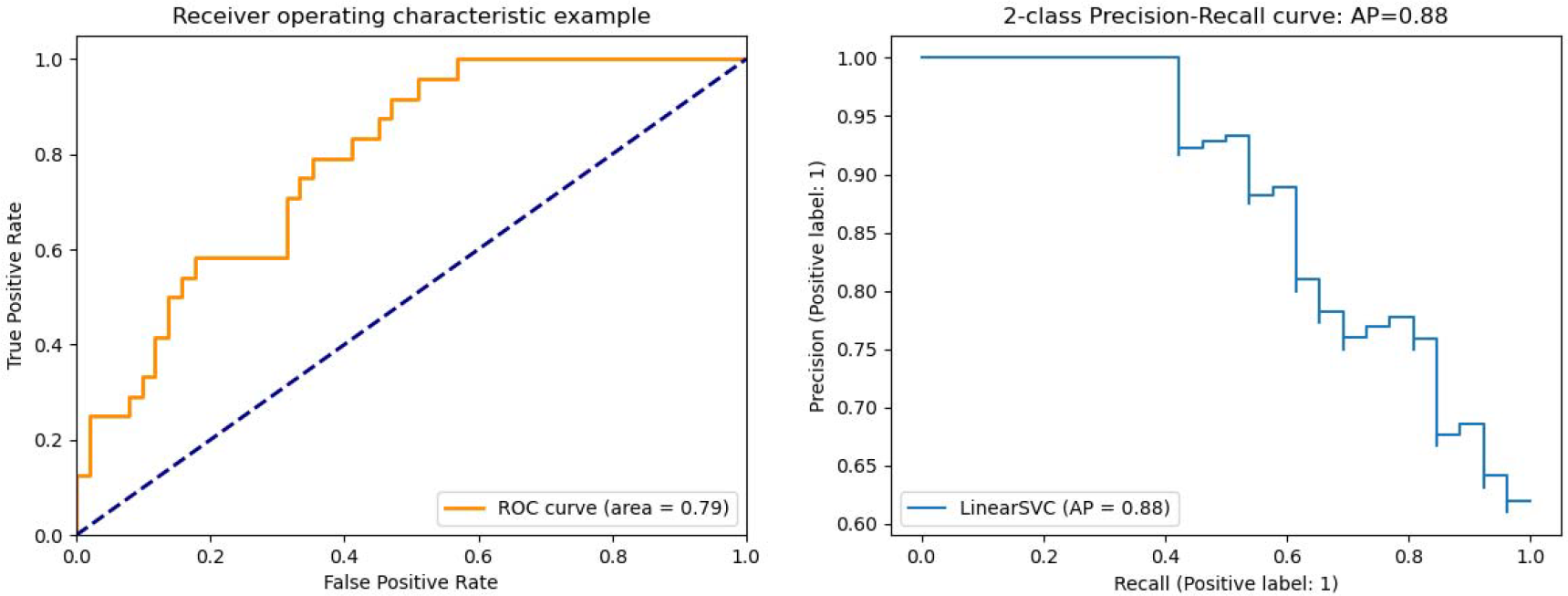
Example of ROC curves and *Precision-Recall*. Reference: ^25^

The ROC Curve allows us to observe how the distribution between the True Positive Ratio (TPR) and the False Positive Ratio (FPR) occurs for different *thresholds*, defined between 0 and 1.

The calculation of both is defined by:

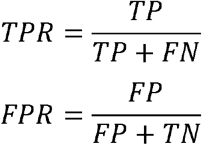

The same principle is present in the curve *Precision* x *Recall* curve.

## 3. Results and Discussion

### 3.1. Correlations

#### Main correlations of the variables in the questionnaire for Diagnosis of Neuropathic Pain 4 (DN4)

Our results in Figure 2 present the main correlations with the variables in the Questionnaire for Diagnosis of Neuropathic Pain 4. It can be seen that 79.19% of the patients who indicated feeling the symptom of numbness in the region where they feel pain also noted that the pain has the characteristic of burning. Notably, 68.6% of people who said they had the burning symptom in the region where they feel pain marked the option of feeling back pain.

**Figure 2.**
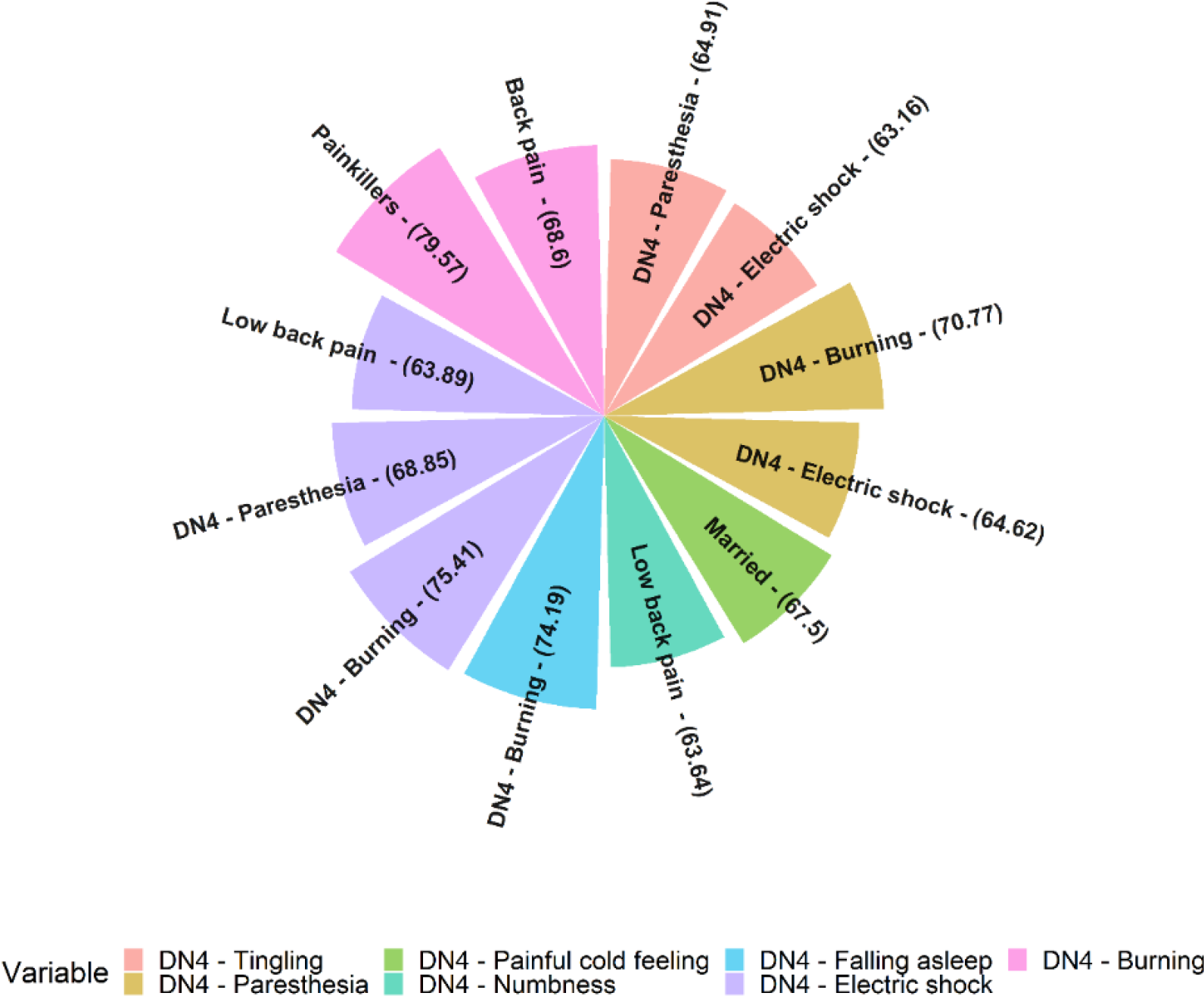
Main correlations of the variables in the Questionnaire for Diagnosis of Neuropathic Pain 4 (DN4).

Figure 3 presents the questions with ordinal variables with the highest correlation with the binary question variables, whether or not the patient presents paresthesia, that is, feeling “pins and needles” at the pain’s region.

**Figure 3.**
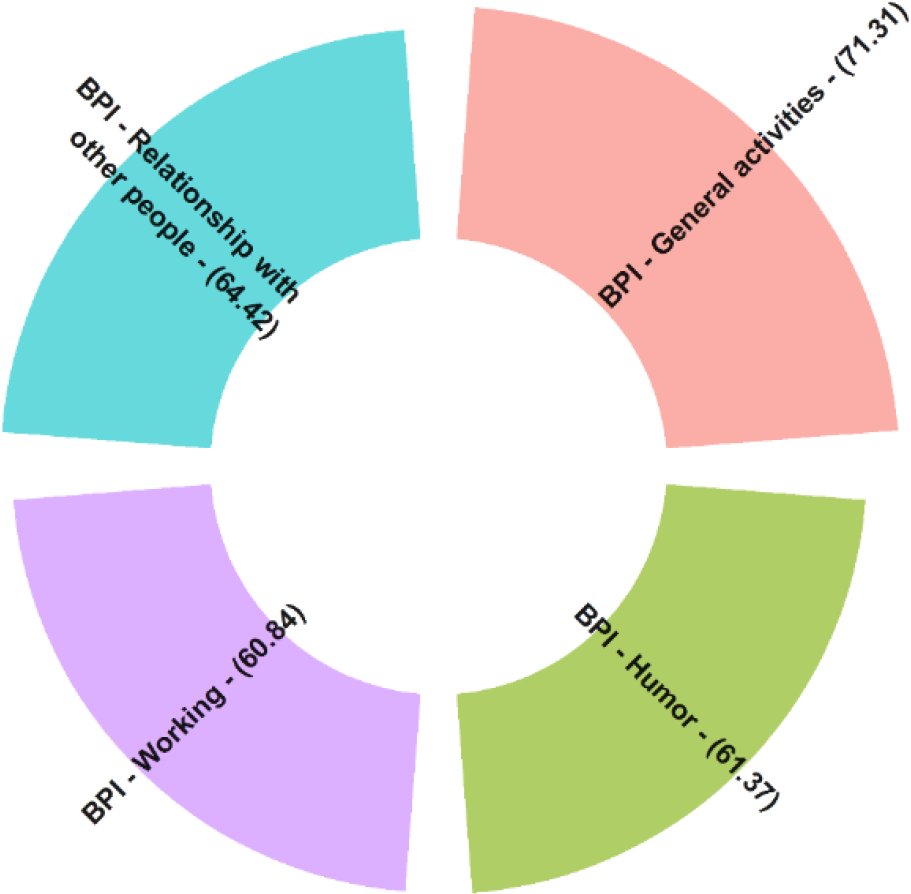
Main correlations between the Questionnaire DN4 about whether or not the patient presents parenthesia - feeling “pins and needles” - at the pain’s region.

Figure 4 demonstrates that patients who reported paresthesia in the pain region also exhibited higher interference of this pain in their relationships with other people, as evidenced by the dense distribution of points and the yellow boxplot representing option 1 of the binary question, which explains the correlation value of 64.42%.

**Figure 4.**
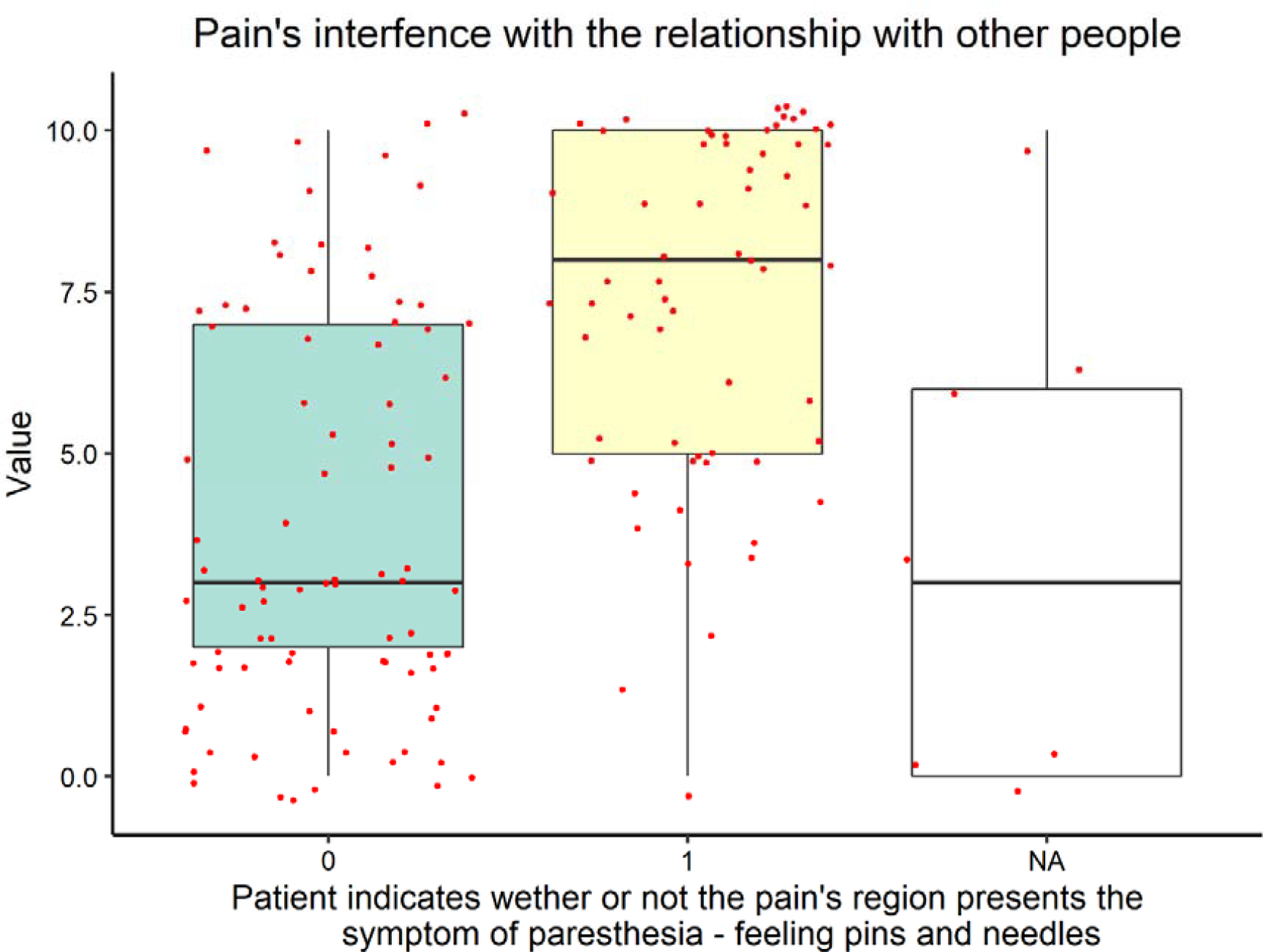
*Boxplot* graph to visualize the data distribution in question 2-b of the DN4 questionnaire with the interference of pain in the relationship with other people.

Notably, only this question (2 - b - pins and needles) from the questionnaire for diagnosing neuropathic pain presented more than 30 people who answered yes or no to the question. The other questions in the DN4 questionnaire did not present statistical significance for the Point Biserial Correlation Coefficient or presented unbalanced data between yes or no; that is, a much more significant proportion of people answered the yes or no option compared to the other alternative.

#### Main correlations between the variables low back pain, leg pain, and back pain with the questions presented in the questionnaires Oswestry 2.0 Disability and Quality of Life Index SF-12

##### Variables of ordinal character

Table 3 contains the main ordinal variables correlated with low back pain from the Oswestry 2.0 disability and quality of life Questionnaire SF-12.

**Table 3.** Ordinal variables of the Oswestry 2.0 disability and quality of life questionnaires SF-12 strongly correlated with low back pain.

| Ordinal | Correlation<br>coefficient | P-value |
| --- | --- | --- |
| SF12_M1_1_v1 | 0.43 | 6.63E-05 |
| Oswestry1_v1 | 0.38 | 5.46E-04 |
| Oswestry2_v1 | 0.37 | 6.07E-04 |
| SF12_M1_8_v1 | 0.35 | 1.25E-03 |
| Oswestry9_v1 | 0.32 | 3.33E-03 |
| Oswestry4_v1 | 0.31 | 4.13E-03 |
| Sum_Oswestry | 0.31 | 4.78E-03 |
| Oswestry10_v1 | 0.31 | 5.08E-03 |
| SF12_M1_10_v1 | -0.30 | 5.92E-03 |
| SF12_M1_12_v1 | -0.36 | 1.07E-03 |
| SF12_M1_11_v1 | -0.37 | 7.40E-04 |
| SF12_M1_9_v1 | -0.38 | 5.77E-04 |
| SF12_M1_2_v1 | -0.40 | 1.97E-04 |
| SF12_M1_5_v1 | -0.46 | 1.53E-05 |
| SF12_M1_6_v1 | -0.47 | 1.07E-05 |
| SF12_M1_3_v1 | -0.48 | 9.29E-06 |
| SF12_M1_7_v1 | -0.49 | 4.68E-06 |
| SF12_M1_4_v1 | -0.51 | 1.59E-06 |

It should be noted that the first question in the SF-12 Quality of Life questionnaire, coded as SF12_M1_1_, presented the highest Bisserial Point Correlation Coefficient, with a value of 0.43. The questions are written in order to enable the patient to inform in a general way about his life quality on a scale that goes from: “weak,” “reasonable,” “good,” “very good,” and “excellent,” listed such as 5,4,3,2 and 1, respectively. Thus, it indicates that patients who reported having low back pain are also more likely to answer this question with the option of life quality closer to “reasonable” or “weak.”

The variable that presented the highest value in the module was the one correlated to the question coded as SF12-M1_4_v1 in the value of -0.51, which asks the patient, in the last four weeks, they ended up doing less than they wanted as a result of their physical state in daily activities at work. The alternatives consist of a scale that goes from “always,” “most of the time,” “some time,” “little time,” and “never,” listed as 1,2,3,4 and 5, respectively. This result indicates that patients with low back pain are more likely to indicate that they “always” or “most of the time” performed less than they wanted at work or other regular daily activities due to their physical condition.

Furthermore, the correlation regarding the low back pain condition with the question SF12_M1_3_v1 presented a value of -0·48, which indicates that the patient’s current health limits him in the activity of climbing several stairways and also with the question SF12_M1_6_v1 (correlation value of -0.47), regarding how much the patient had emotional problems that led him to perform less than he wanted in regular daily activities.

It is also complemented that the question coded as Oswestry1_v1 measures the intensity of the patient’s pain at the moment on a scale ranging from 1 to 6. This variable had a correlation coefficient of 0.38, indicating that patients with low back pain are likelier to have more severe pain. Table 4 brings the two issues most correlated with the issue of leg pain.

**Table 4.**
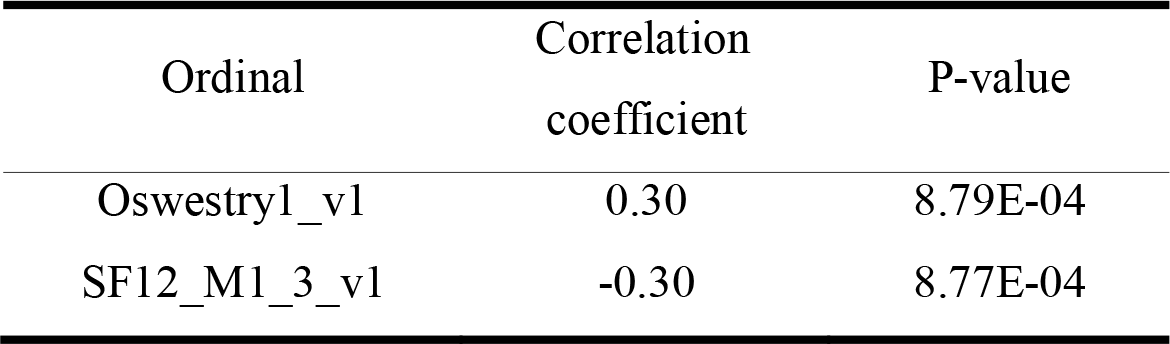
Ordinal variables from the RMDQ, Oswestry, and SF-12 questionnaires strongly correlated with leg pain.

| Ordinal | Correlation coefficient | P-value |
| --- | --- | --- |
| Oswestry1_v1 | 0.30 | 8.79E-04 |
| SF12_M1_3_v1 | -0.30 | 8.77E-04 |

The question coded as Oswestry1_v1 asks how much pain the patient currently feels. Thus, patients with leg pain are also more likely to have a positive result for this issue. In addition, the question in the SF-12 Quality of Life questionnaire, coded as SF12-M1-3-v1, related to how much the patient’s health limits him in climbing several flights of stairs, correlates with leg pain.

Table 5 shows which issues correlate most with the general presence of back pain in patients. It appears that the question from the quality of life SF-12 Questionnaire coded as SF12_M1_8_V1, which asks how the pain interfered with the patient’s routine work in the last four weeks, on a scale of: “absolutely nothing,” “a little,” “moderately,” “a lot” and “immensely,” listed as 1,2,3,4 and 5 respectively, are correlated with the presence or absence of back pain in patients. This way, those who answered that they have back pain are more likely to indicate that the pain interfered “a lot” or “immensely” in their routine work in the last four weeks.

**Table 5.**
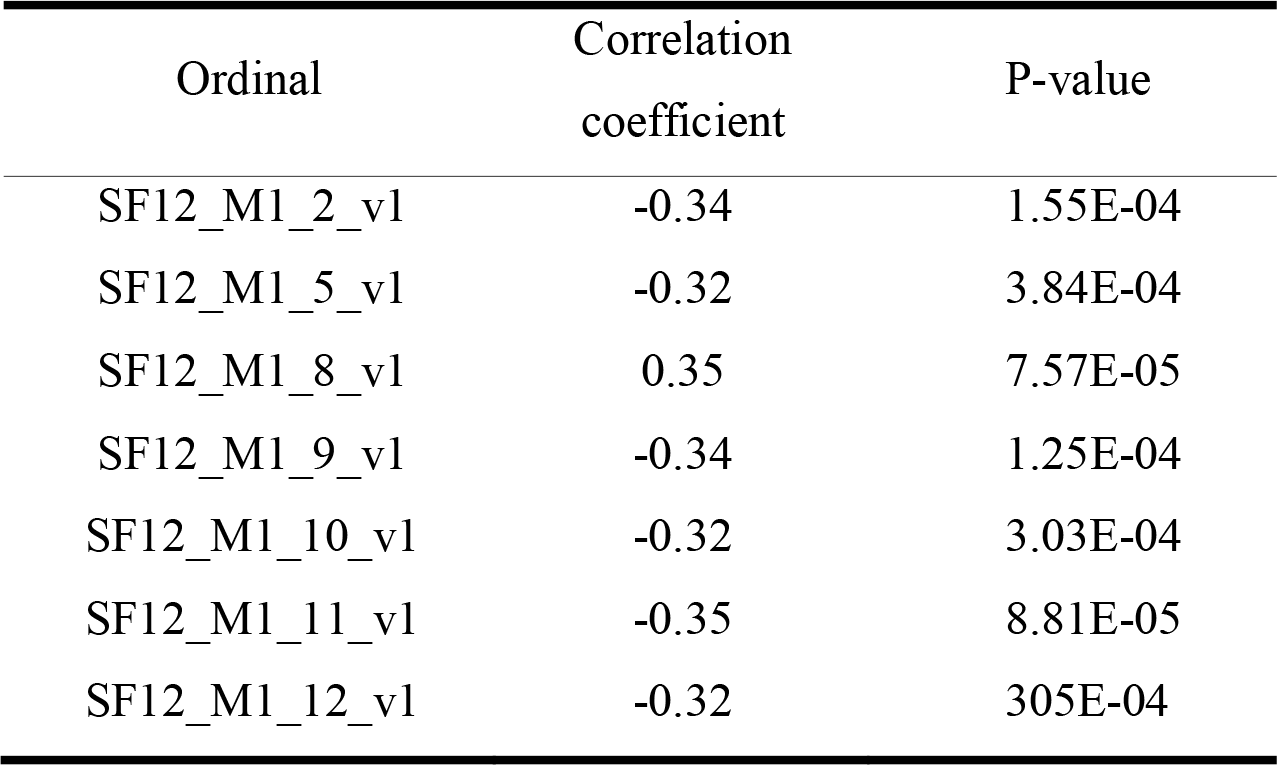
Ordinal variables from the SF-12 Quality of Life questionnaire most strongly correlated with back pain.

The question coded as SF12_M1_11_v1 raises the question of how long, during the last four weeks, the patient felt sad or depressed with the following answers: “always,” “most of the time,” “some time,” “little time” and “never,” listed as 1,2,3,4 and 5. Hence, given the negative correlation coefficient of -0.35, it is concluded that patients with back pain are likelier to answer this question with the options “always” and “most of the time.”

Figure 5 provides a visual summary of the correlations found in Tables 3, 4, and 5 with and previously commented in text form.

**Figure 5.**
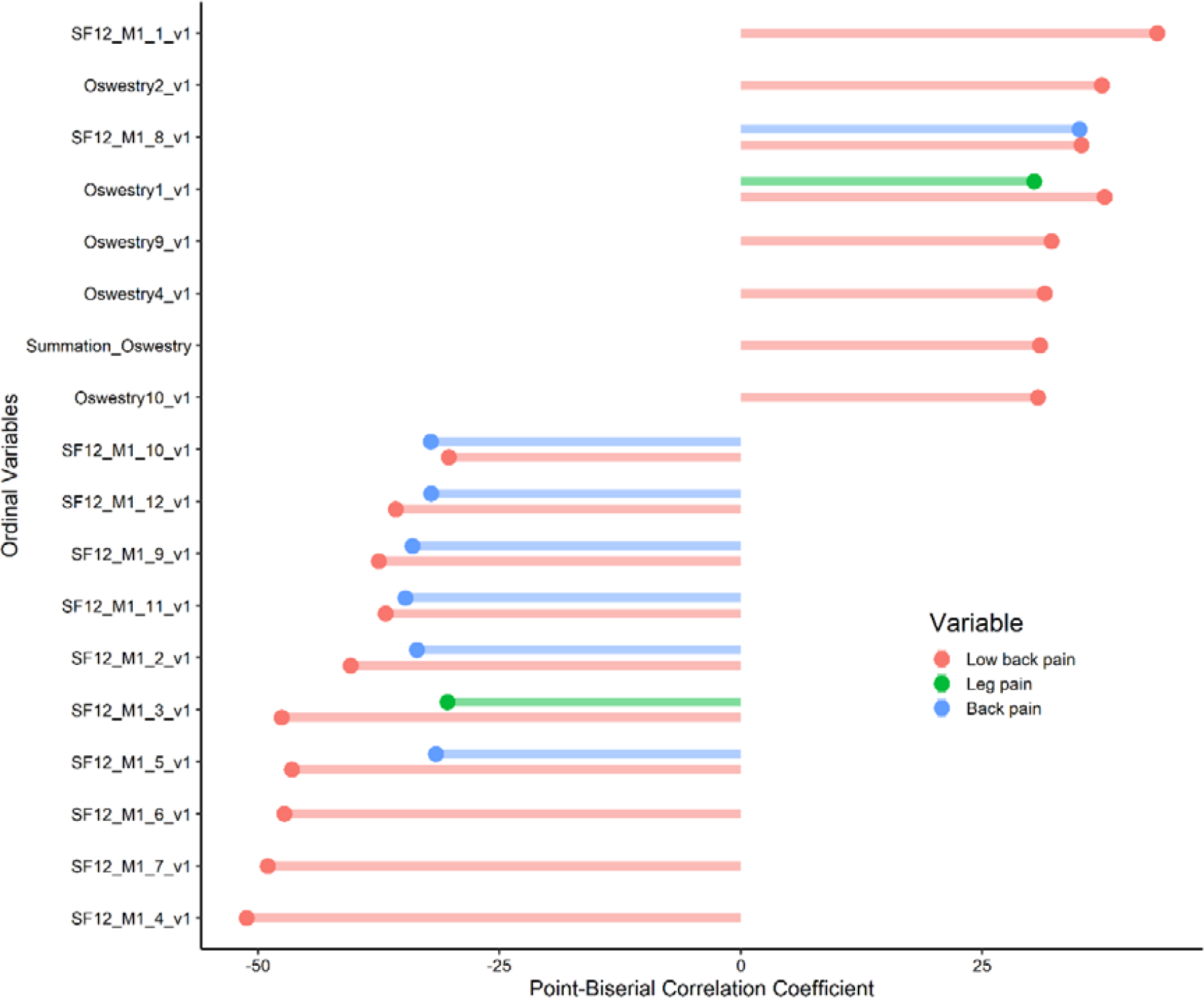
Ordinal questions from the RMDQ, Oswestry, and SF-12 questionnaires most strongly correlated with Low Back Pain, Leg Pain, and Back Pain.

#### Main correlations between the binary variables back pain, low back pain, leg pain, and being unemployed or not

In Figure 6, it is possible to visualize which issues correlate most with the variables low back pain, leg pain, back pain, and whether or not interviewed are unemployed. It is noted that 65.57 % of people who checked the option of presenting pain in the lower back (low back pain) also indicated pain in their legs.

**Figure 6.**
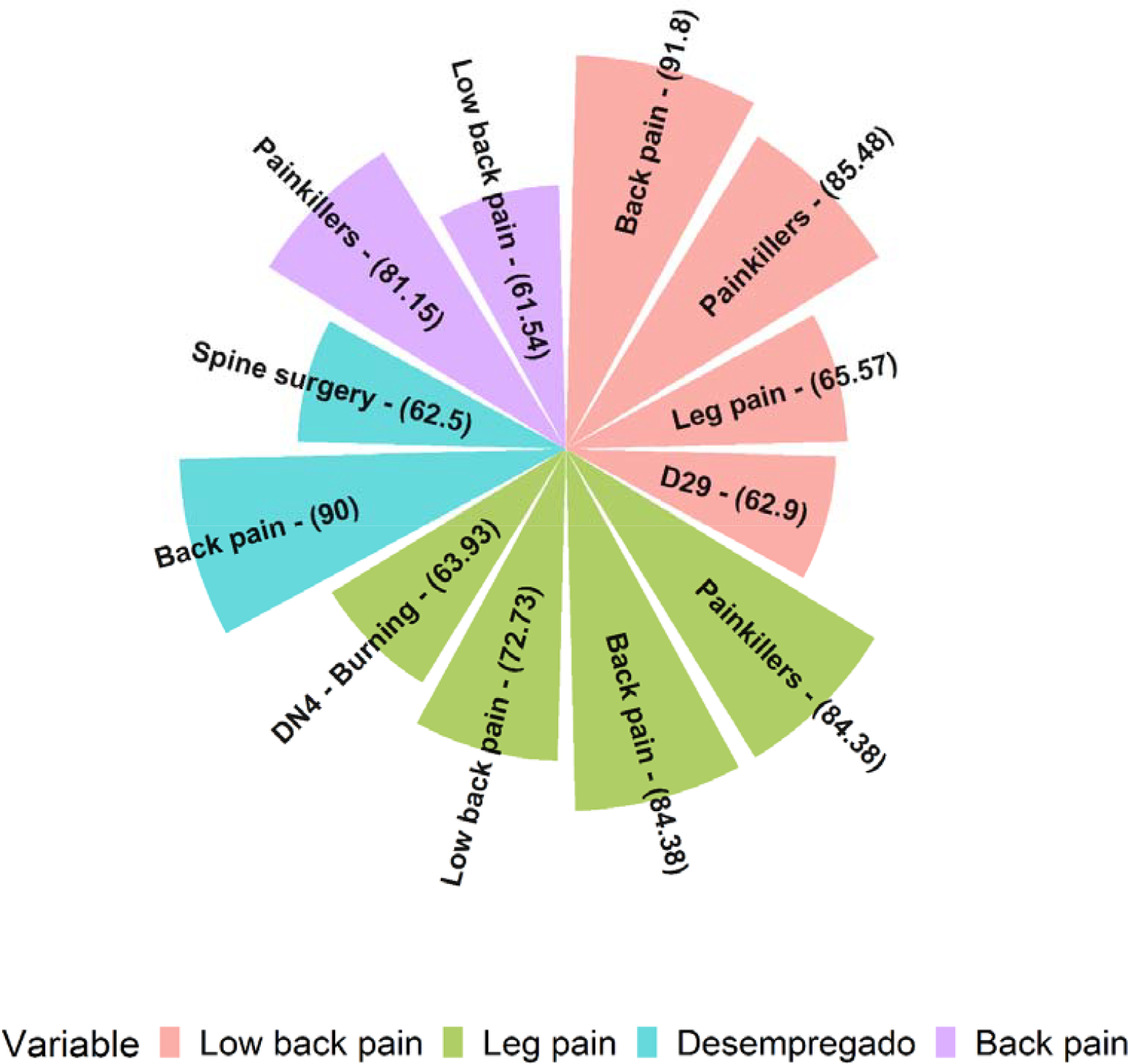
Main correlations between the binary variables back pain, low back pain, leg pain, and being unemployed or not.

It is also important to mention that 63.93 % of patients with leg pain also checked the option in Questionnaire DN4 that their pain has a burning characteristic (DN - burning). The high percentage of patients who use painkillers is also noted, 84.38 % for those with leg pain and 90 % for those with back pain.

Besides that, no correlation was more significant than 60 %, with a significance level of 5 % for the variable that considered whether the patient had already undergone spinal surgery.

### 3.2. Feature Selection

Based on the correlations presented, the following criteria were defined for the selection of predictor variables that are used in the machine learning model:

- According to the contingency table, those variables present a percentage higher than a 60% correlation value;
- From the correlation between the low back pain binary variable (answering yes or no for that question), only those with a correlation more significant than 0·5 or less than -0·5 (values presented in the figures as multiplied by 100).

Thus, the selected variables were:

- Gothenburg - Whether or not you use painkillers;
- Gothenburg - Whether or not you have back pain;
- Gothenburg - Whether or not you have leg pain;
- Gothenburg - Which gender (Male or Female);
- BPI - If you have pain in region 29 of question 2 from the Brief Pain Inventory Questionnaire;
- Gothenburg - How long have you had pain in your legs;
- BPI - Pain intensity at the moment;
- Short Form Health Survey - Performed less than he wanted in daily activities due to pain issues.

### 3.3. Principal component analysis

From the principal component analysis, it is possible to see the Scree Plot graph in Figure 7, which allows us to infer the percentage of variance explained according to the number of main components used. Thus, it is observed that two components can explain up to approximately 59 % of the variability of the data, and with five components, it explains up to 94 %.

**Figure 7.**
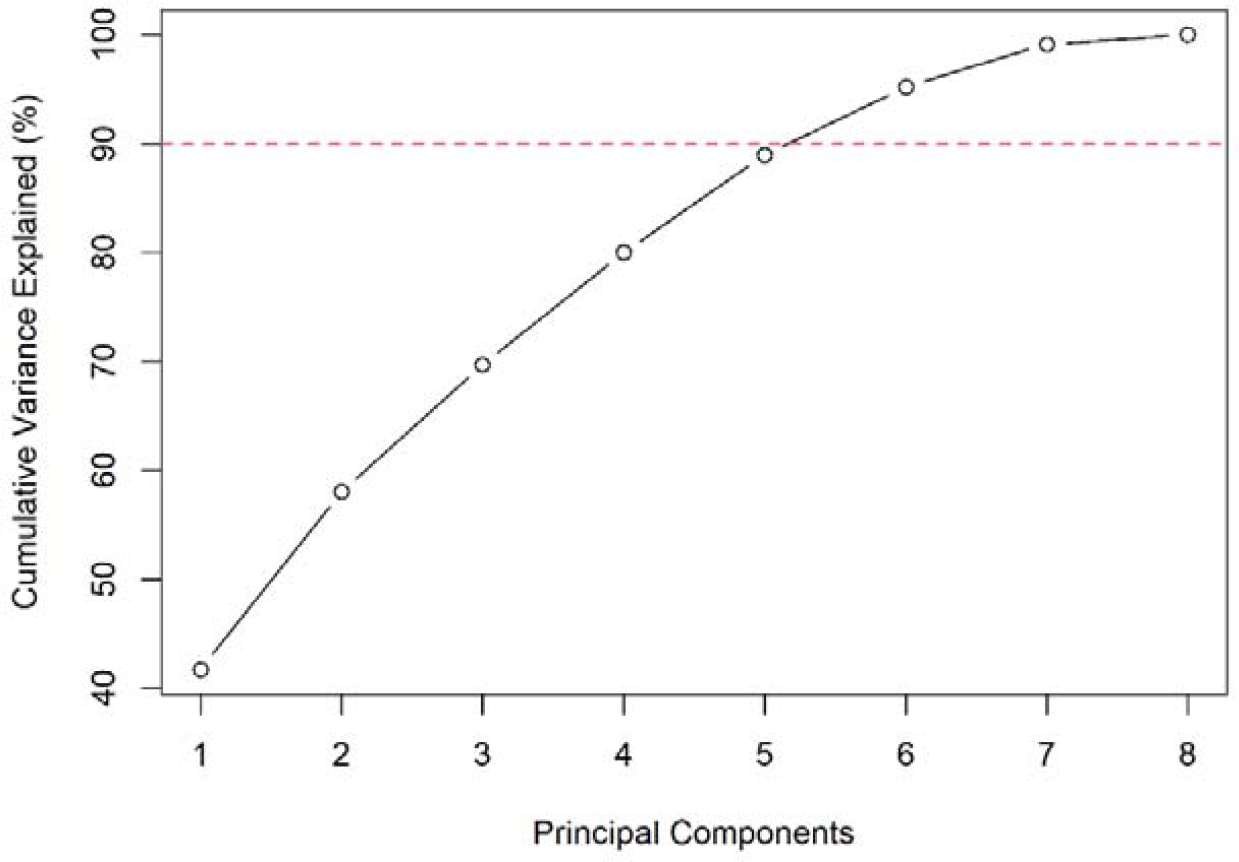
*Scree plot* from the principal component analysis

Since two main components are sufficient to explain up to approximately 59 % of the data variability, it can be seen in Figure 8 how the data are distributed in two dimensions.

**Figure 8.**
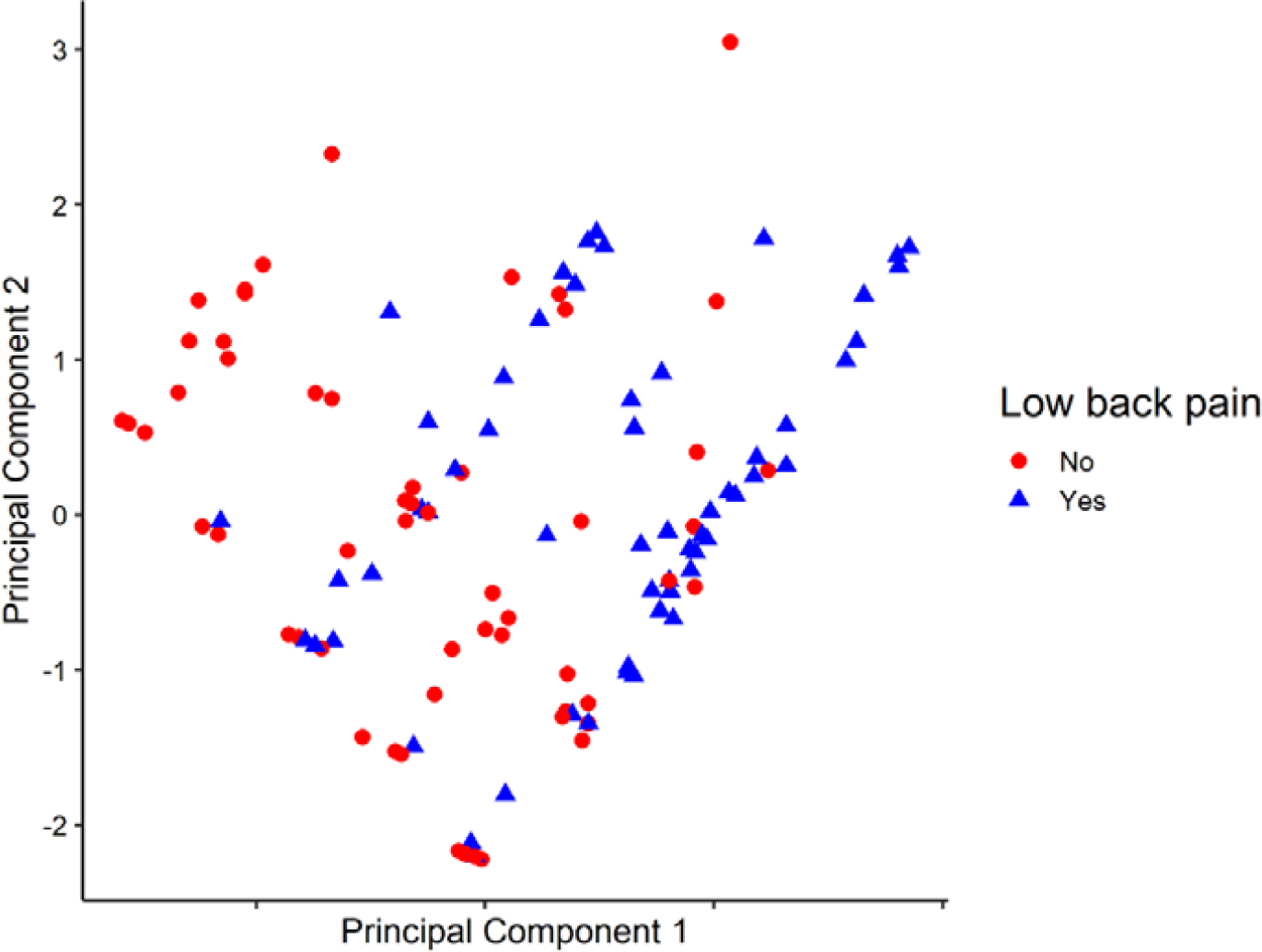
Two-axis view of the distribution of low back pain cases using the first two main components.

It is possible to observe from Figure 8 that with only two dimensions, there is a great presence of overlap between positive and negative cases.

### 3.4. Machine learning model to predict low back pain

From the application of the five machine learning models previously presented in the Methodological procedure section, Figure 9 shows the result of the ROC curve obtained, and in Figure 10 there is the Precision-Recall curve, considering the variation of the *threshold* for prediction between the values 0 and 1.

**Figure 9.**
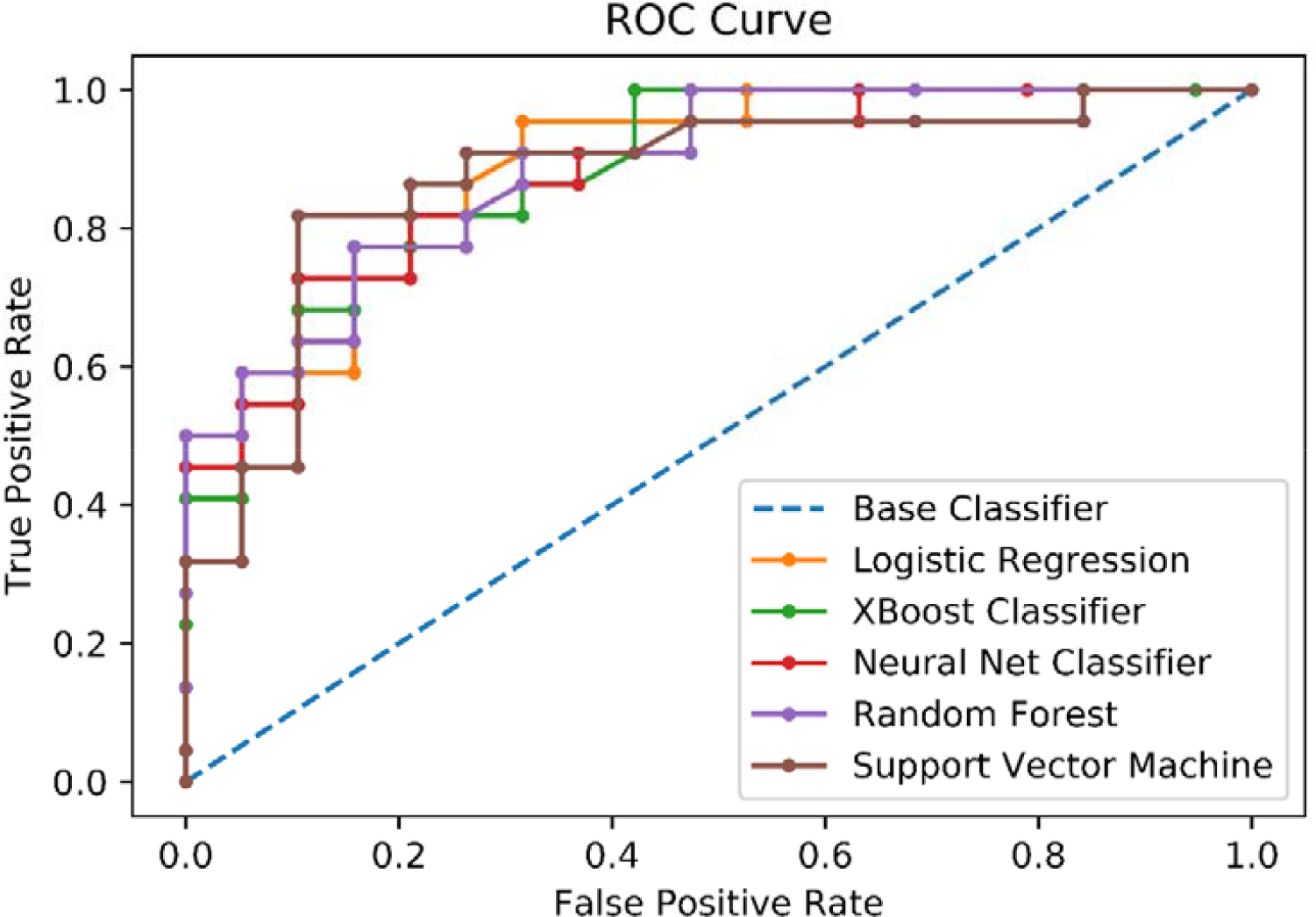
ROC curve for the five different machine learning models.

**Figure 10.**
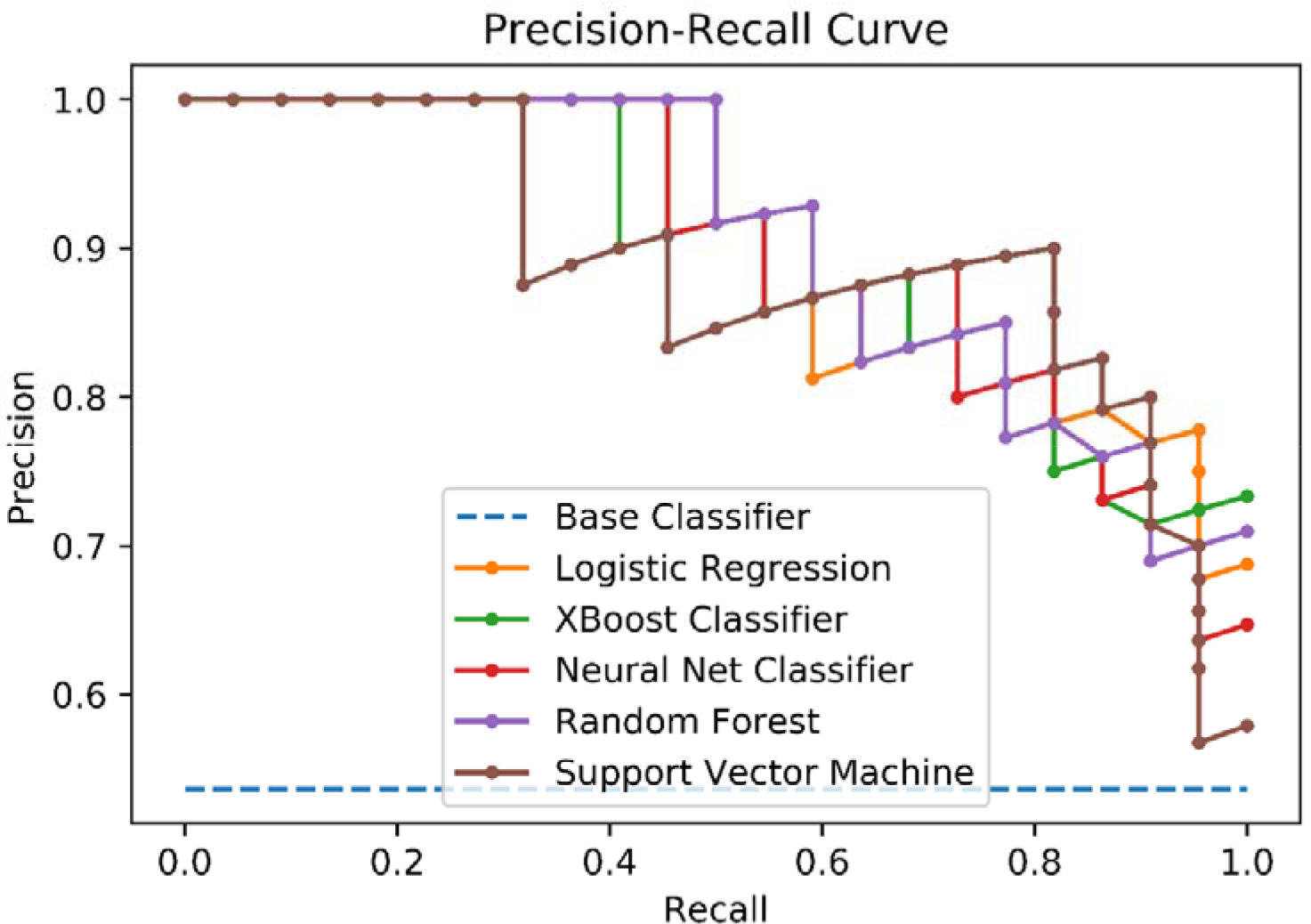
Precision-recall curve for the five different machine learning models.

For the *threshold* default value of 0.5, we present in Table 6 the final metrics obtained with the application of 5 different machine learning models’ performances for classifying patients regarding the presence of low back pain.

**Table 6.** Results of the performance metrics of the tested machine learning models.

| Algorithm | Accuracy | Precision | Recall | F1-Score |
| --- | --- | --- | --- | --- |
| Logistic regression | 0.78 | 0.82 | 0.73 | 0.73 |
| Neural Network | 0.71 | 0.83 | 0.81 | 0.82 |
| Random Forest | 0.78 | 0.77 | 0.74 | 0.74 |
| Support Vector Machine | 0.79 | 0.88 | 0.69 | 0.72 |
| XBoost Classifier | 0.81 | 0.81 | 0.80 | 0.79 |

Therefore, it is noted that with the use of the *threshold* standard of 0.5, the algorithm Neural Network presented the highest *Recall* and *F1-Score* value, and the model built from the *XBoost* algorithm presented the highest accuracy value, as well as the second highest value of *Recall* and *F1-Score*. It should be mentioned that the performance of these models can be improved by collecting more extensive databases and with a smaller proportion of missing values.

It is noteworthy that the task performed and exposed, the discovery of which are the main variables with the highest correlation values with low back pain, allows the reduction of the number of questions in the applied questionnaires. Thus, one of the main benefits of adopting this measure is that patients are more likely to answer questionnaires with fewer questions than the current ones, thus contributing positively to a better quality database generation and data integrity, consequently assisting the patient screening process.

### 3.5. Final considerations

Our findings show a strong link between numbness sensations and burning pain. Furthermore, we discovered a link between burning symptoms and back pain, which impacted patients’ interpersonal connections. Individuals suffering from low back discomfort also had a lower quality of life. They reported limits in work responsibilities and normal activities due to their physical condition, which affected not only their low back pain but also their back pain and leg discomfort. Emotional issues that interfere with regular activities were also addressed. Furthermore, those with low back pain were more likely to have severe pain.

Regarding machine learning, the Neural Network had the highest Recall and F1-Score, while the XBoost approach had the most accuracy and the second highest Recall and F1-Score. It is worth noting that enhancing model performance can be achieved by collecting more extensive databases and minimizing missing variables.

Another example from the literature is a study by Depintor et al. (2016) investigating chronic spinal pain using statistical methods ^25^. The authors utilized Cox Regression (Proportional Risks model) and conducted a bivariate analysis using the Log-Rank test to determine statistical associations. The study, involving 826 participants, estimated the prevalence of chronic vertebral pain at 22% with a confidence interval of 19.3% - 25% at a significance level of 5.0%. Factors associated with chronic vertebral pain included being female, aged 30 or older, having four years or less of schooling, exhibiting symptoms compatible with anxiety, and engaging in intense physical effort during the main occupation ^25^.

Furthermore, a literature review by Tagliaferri et al. (2020) ^16^ evaluated the application of Machine Learning (ML) algorithms for back and lower back pain. The review selected 48 articles for analysis and compared them with established methods such as STarT Back and McKenzie. Among the selected articles, 45 employed samples smaller than 1000, 19 utilized fewer than five parameters in the final model, 13 applied multiple models with high accuracy, and 25 focused on the binary classification of low back pain (presence or absence of pain) ^16^. These studies corroborated our data using statistical and machine learning tools to enhance the management of chronic pain patients, improving their overall quality of life.

## 4. Conclusions

In summary, the presented method allowed us to determine which questions correlate more with back pain, leg pain, and chronic low back pain. Moreover, the machine learning algorithms made use of only seven variables as input to predict the occurrence of chronic low back pain. The Neural Network and XBoost algorithm resulted in the best metrics performance with Recall and Precision near the value of 0.8. These results are essential to assist in the patient screening and allocation process, contributing to reducing costs, as stated by the value-based health care management system approach. Additionally, reducing the number of questions increases the probability of more patients answering every item in a questionnaire and contributes to building larger datasets that can be used to build models with even better metrics.

## Data Availability

The study was approved by the Ethics Committee for Research Project Analysis of the Hospital, under Opinion number 3.239.039 - CAAE: 10024819.2.0000.5485 (Plataforma Brasil), and all study participants were previously informed about the research with both oral and written explanations of the Informed Consent Form and provided written authorization to participate in this program.

## Consent to participate

Informed consent was obtained from all individual participants included in the study.

## Acknowledgments

Also, we would like to thank the institutions of the Instituto de Pesquisas Tecnológicas (IPT) and the University of São Paulo.

## Authors contribution

VPB and JLMSR co-wrote the first draft, which the entire group (VPB, JLMSR, CZVN, NNPC, and VMPG) revised. NNPC and VMPG co-supervised the work.

## Supplementary Data

There is no appendix or supplementary data in this manuscript.

